# Structural and functional pathology in cocaine use disorder with polysubstance use: a multimodal fusion approach

**DOI:** 10.1101/2023.02.20.23285655

**Authors:** Jalil Rasgado-Toledo, Sai Siddharth Duvvada, Apurva Shah, Madhura Ingalhalikar, Vinoo Alluri, Eduardo A. Garza-Villarreal

**Affiliations:** Instituto de Neurobiología, Universidad Nacional Autónoma de México campus Juriquilla, Querétaro, México; Cognitive Science Lab, Kohli Centre for Intelligent Systems, International Institute of Information Technology, Hyderabad, India

**Keywords:** Cocaine use disorder, Imaging, Graph theory, sMRI, fMRI, Multimodal fusion, mCCA + jICA

## Abstract

Cocaine use disorder (CUD) is described as a compulsive urge to seek and consume cocaine despite the inimical consequences. MRI studies from different modalities have shown that CUD patients exhibit structural and/or functional connectivity pathology among several brain regions. Nevertheless, both connectivities are commonly studied and analyzed separately, which may potentially obscure its relationship between them, and with the clinical pathology. Here, we compare and contrast structural and functional brain networks in CUD patients and healthy controls (HC) using multimodal fusion. The sample consisted of 63 (8 females) CUD patients and 42 (9 females) healthy controls (HC), recruited as part of the SUDMEX CONN database. For this, we computed a battery of graph-based measures from multi-shell diffusion-weighted imaging and resting state fc-fMRI to quantify local and global connectivity. Then we used multimodal canonical component analysis plus joint independent component analysis (mCCA+jICA) to compare between techniques, and evaluate group differences and its association with clinical alteration. Unimodal results showed a striatal decrease in the participation coefficient, but applied supervised data fusion revealed other regions with cocaine-related alterations in joint functional communication. When performing multimodal fusion analysis, we observed a higher centrality of the interrelationship and a lower participation coefficient in patients with CUD. In contrast to the unimodal approach, the multimodal fusion method was able to reveal latent information about brain regions involved in impairment due to cocaine abuse. The present results could help in understanding the pathology of CUD in order to develop better pre-treatment/post-treatment intervention designs.

## Introduction

Cocaine use disorder (CUD) is described as a compulsive urge to seek and consume cocaine despite the inimical consequences. CUD causes a gradual decline of the patient’s cognitive and behavioral health (1) along with greater health and socioeconomic issues. MRI studies from different modalities have shown that CUD patients exhibit structural and/or functional connectivity pathology among a wide variety of regions such as frontal, parietal, temporal gyri, and subcortical regions, including white matter tracts that connect these regions (2,3). A recent meta-analysis showed that CUD patients display lower volume in the orbitofrontal cortex, temporal pole, anterior insula, anterior thalamic radiation, cingulum, inferior occipitofrontal fascicle, and acoustic radiation (4). Some studies also show brain network alterations using graph theory in CUD with non-consistent results (5–7). Nevertheless, structural and functional MRI pathology is commonly studied and analyzed separately, which may potentially obscure the relationship between them, and clinical pathology.

Multimodal fusion provides a means to reveal complicated hidden relationships between modalities and weak latent effects in high-dimensional data by taking advantage of the presence of cross-information in cross-individual variance (8,9). Multimodal fusion has the added benefit of increased robustness to modality-specific noise (9) and has been used to study different brain pathologies like schizophrenia, bipolar and obsessive-compulsive disorders (10–12). A recent study by Meade et al. (13) explored multimodal fusion techniques namely multimodal canonical component analysis (mCCA) in conjunction with joint independent component analysis (jICA) on CUD patients using whole-brain voxel-wise maps and their relation with the delay discounting task. This study showed structural and functional co-alterations in CUD patients, linked to impulsive behavior. The relevance of multimodal fusion techniques can be leveraged to develop models that can exploit the data and minimize incorrect conclusions in psychiatric disorders (8,9).

In the present study, we wished to compare and contrast structural and functional brain networks in CUD patients and healthy controls (HC) using multimodal fusion. Unlike other multimodal fusion studies which commonly use voxel-based metrics, we use graph theory metrics which enable us to meaningfully understand and analyze brain connectivity architecture. Compared to voxel-based analysis, graph-based analysis of the brain offers a better mathematical framework to model the communications between various brain regions. For this, we computed a battery of graph measures from multi-shell diffusion-weighted imaging and resting state functional connectivity to quantify local and global connectivity. Then we used jICA and mCCA+jICA to compare techniques and evaluate group differences and their association with the clinical alteration.

## Methods

### 1. Participants

The sample consisted of 63 (8 females) cocaine use disorder patients (CUD) and 42 (9 females) healthy controls (HC), recruited as part of the SUDMEX CONN database (14), paired by age, sex, handedness, and education. We included participants with T1-weighted, diffusion-weighted imaging (DWI), and resting-state fMRI (rsfMRI) sequences for this study. Due to analysis failures, three CUD patients and one HC subject were eliminated from the analysis. Demographic characteristics are shown in Table 1. According to the Declaration of Helsinki, the ethics committee of the Instituto Nacional de Psiquiatría “Ramón de la Fuente Muñiz’’ in Mexico City, Mexico gave the ethical approval for this work. The entire study was carried out at the same Institute. All participants provided verbal and written informed consent. Recruitment criteria and full sample details are described in Angeles-Valdez (14).

**Table 1.** Demographic characteristics of participants.

|  | CUD (n=63) | HC (n=42) | Stats |
| --- | --- | --- | --- |
| Age | 32 (18 - 50) | 30 (18 - 48) | $t = -0.5, p = 0.6$ |
| Education | Middle School | High School | $\chi^2 = 10, p = 0.04$ |
| Handedness * | (n = 63) | (n = 42) |  |
| Right | 56 | 35 |  |
| Left | 4 | 4 |  |
| Ambidextrous | 3 | 3 |  |
| Onset age of consumption | 20 (12 - 41) | na |  |
| Years consuming | 10 (1 - 28) | na |  |
| Average consumption per intake * | (n = 55) | na |  |
| < 0.8 gm | 0 | na |  |
| 1.6 - 2.4 gm | 7 | na |  |
| 3.2 - 5.6 gm | 11 | na |  |
| 8 - 9 gm | 26 | na |  |
| 9 - 10 gm | 6 | na |  |
| > 10 gm | 10 | na |  |

Median (min – max) for all except: \* = count. CUD = Cocaine Use Disorder, HC = Healthy controls, n = due to absence of data we show each sample size per variable, na = not applicable, gm = grams, Stats = statistics, t = t-value, $\chi^2$ = chi-squared.

### 2. Magnetic Resonance Imaging

MRI sequences were acquired using a Philips Ingenia 3T system (Philips Healthcare, Best, The Netherlands, and Boston, MA, USA) with a 32-channel dS Head coil. Rs-fMRI was acquired using a GE-EPI sequence with the following parameters: TR = 2000, TE = 30.001ms, flip angle = 75°, matrix = 80×80, FOV = 240 mm^2^, voxel size = 3×3×3 mm, number of slices=36, phase encoding direction = AP. T1-weighted (T1w) was acquired using a three-dimensional FFE SENSE sequence, TR = 7, TE = 3.5 ms, FOV = 240mm^2^, matrix = 240×240 mm, number of slices = 180, gap = 0, plane = sagittal, voxel = 1×1×1 mm. Subjects were instructed to keep their eyes open and try not to think about anything in particular, while a fixation cross was presented. We then acquired a spin-echo High Angular Resolution Diffusion Imaging (DWI-HARDI) sequence, TR = 8600 TE = 126.78 ms, FOV = 224, matrix = 112×112 mm, number of slices = 50, gap = 0, plane = axial, voxel = 2 × 2 × 2 mm, directions: 8 = b0, 36 = b-value 1,000 s/mm² and 92 = b-value 3,000 s/mm², total = 136 directions. The MRI order of acquisition was: 1) rs-MRI, 2) T1w, and 3) DWI-HARDI. The total scanning time lasted around 50 minutes.

### 3. Clinical measures

Participants were evaluated using a battery of paper-based clinical questionnaires before MRI scanning. For this study, the CUD group was assessed using the Cocaine Craving Questionnaire (CCQ-General) and (CCQ-Now) to rate their craving over the previous week and at the time of MRI scanning. This questionnaire includes questions about the desire to use cocaine, the anticipation of positive outcomes, and relief from withdrawal (15). For evaluating functional impairments or disabilities in psychiatric patients, the CUD group was assessed by World Health Organization Disability Assessment Schedule 2.0 (WHODAS 2.0). This instrument has been widely used in different countries for health and disabilities, finding high consistency rates (16). The CUD group was also assessed by Addiction Severity Index (ASIP) for consumption status and addiction severity. This instrument is a semi-structured interview that evaluates several functional domains like medical status, employment, alcohol use, drugs use, family/social life, and psychiatric status (17). Impulsivity was also assessed using the Barratt Impulsiveness Scale Version 11 (BIS-11), which is a self-report scale that assesses three categories of personality/behavioral impulsivity: cognitive (i.e. inability to focus attention), motor (i.e. act without thinking), and non-planning impulsiveness (i.e. lack of forethought) (18). For information on other clinical measures that were recorded along with the above-mentioned, see Angeles-Valdez et al (14).

### 4. Preprocessing

Data preprocessing of rsfMRI and T1w images were performed using fMRIprep pipeline (19). Structural T1 steps included a volume correction of intensity non-uniformity, skull-stripped, brain tissue segmentation, and a spatial normalization onto MNI common brain space (MNI152NLin2009cAsym). Functional preprocessing steps included correction for intensity, slice-timing and head motion, spatial smoothing with an isotropic Gaussian kernel of 6 mm full width at half maximum (FWHM), framewise displacement threshold of 0.5, distortion estimation using a field map, skull-stripped, and a spatial normalization to MNI brain space. Resting-state time series were then processed using XCP Engine v. 1.2.1 (20) with nuisance regression using the pipeline described in Power et al. (21). Shortly, nuisance strategy included: 1) inhomogeneities correction, 2) dummies removal (4 initial volumes), 3) realignment of all volumes to reference, 4) demeaning and removal of trends, 5) co-registration, 6) removal of global, white matter and cerebrospinal confounding signals, 7) motion scrubbing and 8) temporal filtering with a first-order Butterworth filter using a bandpass between 0.01 and 0.08 Hz. DWI-MRI preprocessing consisted of the correction of eddy current artifacts and motion noise using eddy_correct from FSL 6.0.11 (https://fsl.fmrib.ox.ac.uk). Gradients were rotated to match the affine transformation applied in eddy step, subsequently, skull stripping was performed using the brain extraction tool from FSL. Finally, using Advanced Normalization Tools (ANTs) registration, MNI brain space was transformed into individual subject space to map all the ROI into subject space.

### 5. Connectivity Analyses

#### a. Structural Connectome

The diffusion connectivity (tc-dmri) matrices were computed using the Harvard-Oxford (HO) cortical and subcortical atlases (112 regions) and with the Desikan Killiany (DK, 86 regions) atlas through fiber-counting. For T1w structural, data were processed using Freesurfer, which performed intensity bias correction, non-brain tissue discard, and tissue segmentation before performing entire brain parcellation using spherical transformation and surface-based registration with the atlas. Connectivity maps were constructed by whole-brain streamline fiber tractography on native space using MRtrix v.3 (22). All ROIs in each atlas in sequence 1 to n-regions and the probabilistic connectome values represent the connectivity fiber counts between the source and destination ROIs. All fibers that start or end in GM-ROIs were taken into the fiber count. Normalization of all values was performed by summing the number of voxels from the source and destination ROIs. The main analysis presented here is based on the Harvard-Oxford atlas. The analysis using the Desikan Killiany atlas is presented in the Supplementary material with no group significant differences.

#### b. Functional Connectome

The functional connectivity (FC) matrices, or fc-fMRI, were computed using the HO cortical and subcortical atlases and the DK atlas. The mean time series was extracted from each region and the functional connectivity matrix was estimated by computing pairwise Pearson correlations. Following this, the FC matrices were thresholded to generate a binary adjacency matrix that represents the presence or absence of functional connectivity. The thresholding and binarization procedures help reduce weaker connections and result in undirected, unweighted, binary matrices where the correlations above a certain threshold are represented by 1 and 0 otherwise. Since the choice of threshold can be arbitrary, we generated several binarized adjacency matrices by varying the cut-off to include the top 5% to 50% with increments of 5%. The main analysis presented here is based on the Harvard-Oxford atlas. The analysis using the DK atlas is presented in the Supplemental Material with no group significant differences.

#### c. Graph-based measures

Graph theory analyses are performed on the binarized adjacency matrices using Matlab v. 2019b and the Graph Theoretical Network Analysis (GRETNA) toolbox (23). The computed graph measures were classified into two categories based on the type of connectivity they signify: global and local graph measures. These graph measures enable us to understand properties like connectivity or topology, at whole-brain and region levels respectively. Global measures comprise Assortativity (r), Network efficiency (*E*_*global*_), Modularity (M), Smallworld index (σ), and Hierarchy (β). Local measures include Betweenness Centrality (BC), Degree Centrality (DC), Participation Coefficient (PC), Nodal Local Efficiency (NLE), and Nodal clustering Coefficient (NCC). These measures are discussed in supplementary material.

Assortativity is the correlation between the degree of a node and the average degree of the node’s neighbors. Network efficiency is defined as the average inverse shortest path length in a network. Modularity (M) is a statistic used to distinguish between the number of intra-module connections of an existing network and randomly connected edges in a random network and is computed based on a greedy agglomerative method (24). Small world index is defined as the ratio of the normalized clustering coefficient and normalized characteristic path length. The Betweenness Centrality of a node (v) is defined as the ratio of the number of shortest paths passing through the node between any two given nodes σ_*ab*_(*v*) to the total number of shortest paths between the two given nodes σ_*ab*_. The Degree Centrality of a node(v) is defined as the ratio of the degree of the node (*d*_*v*_) to the maximum possible degree of the node. The Participation coefficient of a node(v) reflects the within-module and intermodular communication. Nodal local efficiency is similar to network efficiency, but it is computed in the neighborhood of a node. The clustering coefficient of a node(v) is defined as the ratio of the number of connections between the neighbors of the node (*e*_*v*_) to the total number of possible connections among (*K*_*v*_) neighbors of the node.

Those graph measures also quantify network segregation (Nodal clustering coefficient, nodal local efficiency, small world index, and modularity), network integration (Global efficiency, assortativity, and participation coefficient) and node centrality (Betweenness Centrality and Degree centrality) (25).

### 6. Unimodal Analysis

This was performed to investigate brain network differences from the perspective of each modality (fc-fMRI and tc-dMRI). Global and local graph measures were analyzed to understand the functional and structural connectivity of the different brain regions. The global and local graph measures within each modality were tested for differences between CUD and HC using the Mann-Whitney-Wilcoxon test for independent two-samples. Spearman correlation between the graph measures and their corresponding clinical scores within each modality was performed. To account for multiple comparisons, we used FDR at 5% via the Benjamini-Hochberg procedure. R programming language version 4.1 was used for the statistical analysis.

### 7. Multimodal Fusion

Sui et al, (8) outlined several multimodal fusion methods and divided them into three different categories based on their objectives: finding flexible connections between modalities, separating sources and discovering the common mixing profiles, and examining both flexible modality connections and distinct sources. The several outlined blind source separation methods were evaluated by simulating their performances on a generated dataset of 100 noisy (random Gaussian noise was added) images with fMRI and EEG signals as the two modalities. Based on various metrics, it was evident that jICA and mCCA are good at source separation and modality associations respectively. Whereas, mCCA+jICA had more reliability in estimating the modality relations (high or low correlations) along with good source separation and robustness to noise. Hence, our multimodal fusion analysis was performed using mCCA+jICA. Figure 2 depicts the pipeline of our multimodal fusion analysis. Age was regressed out from the local graph-based measures calculated for each modality. The minimum description length (MDL) method (26) was used to determine the number of independent components (M) to preserve for each local graph metric (M_BC_=4, M_DC_=10, M_NCC_=13, M_NLE_=19, M_PC_=9) in both fc-fMRI and tc-dMRI modalities. Next, mCCA analysis was performed on fc-fMRI and tc-dMRI to produce canonical variates (CV_fc-fMRI_ and CV_tc-dMRI_) and canonical components (CC_fc-fMRI_ and CC_tc-dMRI_) .CC_fc-fMRI_ and CC_tc-dMRI_ were concatenated and subjected to joint ICA, yielding mixing profiles (*MM*), unmixing profiles (*umm*), stability indices (IQ), and independent component loadings (IC). MATLAB v. 2019b was used to calculate the mCCA and jICA analyses with custom scripts and the ICASSO toolbox (27). Effective mixing profiles were calculated for group comparisons using the following equations:

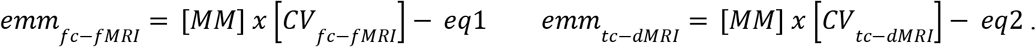

**Figure 1.**
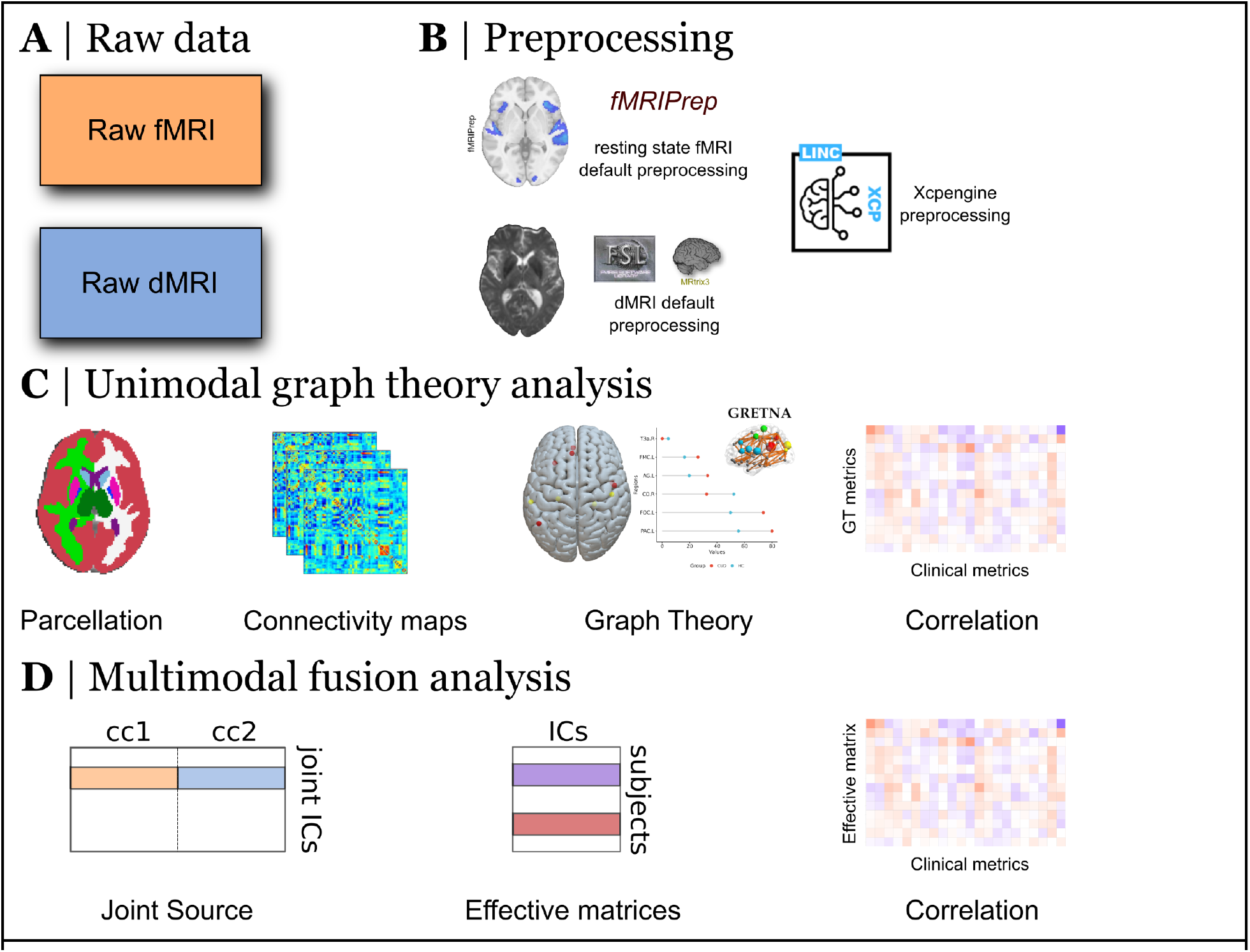
Workflow of all stages analysis. A) Data from 66 (8 females) cocaine use disorder patients (CUD) and 43 (9 females) healthy controls (HC), B) Data preprocessing of Resting-state and T1-weighted MRI images were performed using fMRIprep pipeline follow by XCPengine pipeline, and FSL DWI preprocessing, C) Connectivity analysis based on Graph-based approach along with correlation with clinical measures, D) mCCA+jICA multimodal fusion based on local graph-based measures.

**Figure 2.**
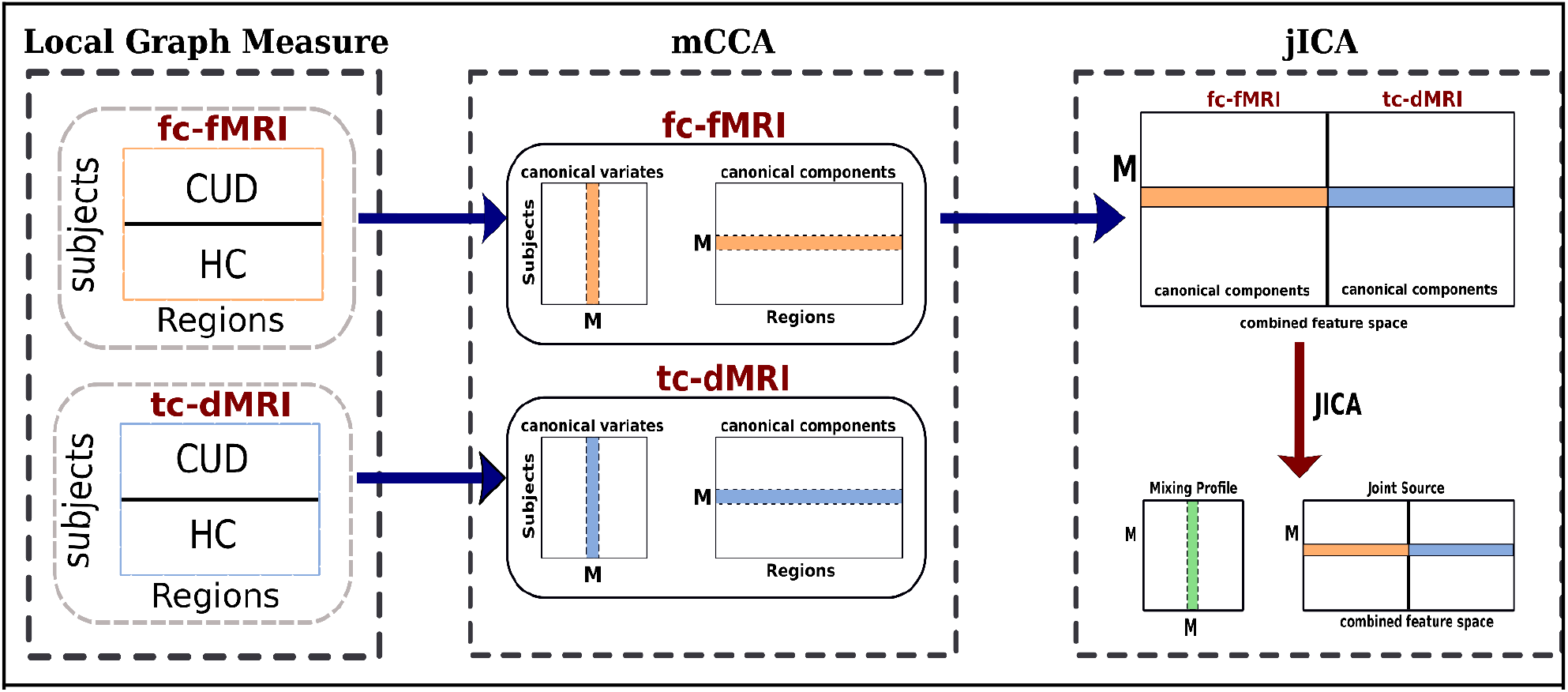
Workflow of the 2-way multimodal fusion analysis using mCCA + jICA fusion strategy of each Local graph measure.

All ICs values were normalized to Z-scores. In order to investigate regions with the highest contribution for the mixing profiles we used a threshold of *Z* ≥ ± 2. 3. The effective mixing profiles (*emm*_*fc−fMRI*_ *and emm*_*tc−dMRI*_) of those ICs with an IQ > 0.8 were considered. Based on the previous research (28), these mixing profiles were further correlated with each other along with their corresponding clinical measures using Spearman correlation. Mann–Whitney–Wilcoxon test for independent two-samples was computed to reveal group differences between CUD and HC for each *emm*_*n*_. All results were corrected for multiple comparisons using FDR (Benjamini-Hochberg) at 5%.

## Results

The present results were generated from the graph measures computed using Harvard Oxford Atlas (HOA), which is known to capture structure–function relationship better than other atlases (29).

### 1. Unimodal Results

No significant differences were observed between CUD and HC groups for the global measures in the tc-dMRI modality. Only the left Caudate exhibited a significant decrease in PC among CUD compared to HC in the fc-fMRI modality (U =1880, p=0.0027, p_fdr_=0.030) (Fig. 3a). For either modality, there were no significant correlations between either the global or local measures and clinical measures.

**Figure 3.**
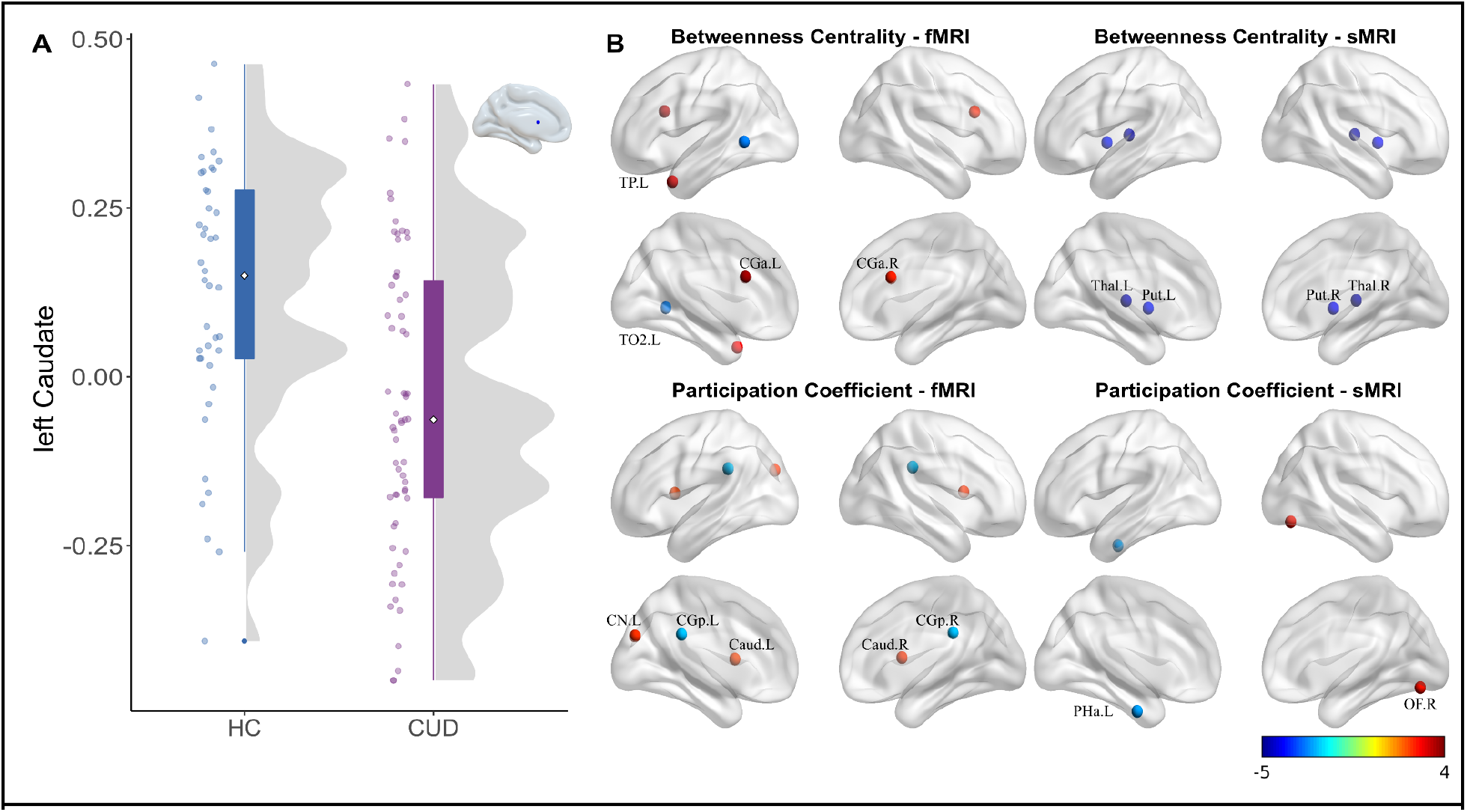
A) Group comparison of Participation Coefficient from unimodal analysis, significant on left Caudate. B) Multimodal group-discriminating *BC − IC*_2_ and *PC − IC*_4_. Each IC component was normalized to z-scores and thresholded to *Z* ≥ ± 2. 3. For Betweenness Centrality in both modalities, positive values means that CUD has higher BC in these regions compared to HC, whereas negative values means the opposite. For the Participation Coefficient in fc-fMRI, positive values mean higher PC in HC than in CUD, whereas negative values mean the opposite. For the Participation Coefficient in dMRI, positive values mean lower PC in HC than in CUD, whereas negative values mean the opposite. TP.L = left Temporal Pole, TO2.L = right middle Temporal gyrus, temporo-occipital, CGa.L = left Cingulate Gyrus, anterior division, CGa.R = right Cingulate Gyrus, anterior division, Thal.L = left Thalamus, Thal.R = right Thalamus, Put.L = left Putamen, Put.R = right Putamen, CGp.L = left Cingulate Gyrus, posterior division, CGp.R = right Cingulate Gyrus, posterior division, CN.L = left Cuneal Cortex, Caud.L = left Caudate, Caud.R = right Caudate, PHa.L = left Parahippocampal Gyrus, anterior division, OF.R = right Occipital Fusiform Gyrus.

### 2. Multimodal Results

The local graph measures of integration, betweenness centrality (BC), and participation coefficient (PC), had IQ > 0.8, resulting in one IC in the case of BC (*IC*_2_) and two for PC (*IC*_1_ and *IC*_4_). Only the fc-fMRI modality showed significant group differences, with BC (*p*_*value*_ = 0. 028, *p*_*fdr*_ = 0. 042) having greater values in the CUD sample and PC (*p*_*value*_ = 0. 026, *p*_*fdr*_ = 0. 042) having lower values in the CUD group. In fc-fMRI modality, the significance of the BC (IC_2_) suggests that the independent component was expressed more strongly in the CUD group compared to the HC group, thus resulting in a lesser number of shortest pathways via nodes (Table 2). For PC, the independent component (IC_4_) that corresponds to the fc-fMRI modality was stronger in the CUD group than the HC, suggesting a higher level of node participation in their own communities (Table 2). The joint-IC_2_ of BC was characterized by the contributions of the left Temporal pole, left Middle Temporal gyrus and bilateral Anterior Cingulate gyrus in fc-fMRI modality along with bilateral Thalamus and bilateral Putamen in tc-dMRI modality (Table 3). In the tc-dMRI modality, the joint-IC_4_ of PC revealed significant connectivity in the left Parahippocampal gyrus and the right Occipital Fusiform gyrus, as well as the bilateral Posterior Cingulate gyrus, left Cuneal cortex, and bilateral Caudate in the fc-fMRI modality (Figure 3B and Table 3). Figure 4 depicts a positive correlation between the effective mixing profiles (*emm*_*fc−fMRI*_ *and emm*_*tc−dMRI*_) of joint IC_2_ and joint IC_4_ for BC and PC, respectively. This demonstrates a strong association between the regions that contribute to the joint independent component in both the fc-fMRI and tc-dMRI modalities for BC and PC. There were no significant correlations with the clinical measures.

**Table 2.** Group comparison in *IC*_*n*_ loadings (IQ > 0.8), median effective mixing profiles of CUD and HC and statistics of each graph metric.

| | | $IC$ | HC (n=42) | CUD (n=63) | Statistic | $p$ value | $p_{fdr}$ |
| --- | --- | --- | --- | --- | --- | --- | --- |
| Betweenness Centrality | fc-fMRI | $IC_2$ | -0.0094 | 0.0088 | $W = 987$ | 0.028* | 0.042* |
| | tc-dMRI | $IC_2$ | 0.0006 | -0.0001 | $W = 1325$ | 0.992 | 0.992 |
| Participation Coefficient | fc-fMRI | $IC_1$ | -0.0111 | -0.0112 | $W = 1352$ | 0.852 | 0.852 |
| | | $IC_4$ | 0.0104 | 0.0027 | $W = 1664$ | 0.026* | 0.042 * |
| | tc-dMRI | $IC_1$ | -0.0038 | 0.0018 | $W = 1143$ | 0.24 | 0.720 |
| | | $IC_4$ | -0.0015 | -0.0011 | $W = 1235$ | 0.567 | 0.850 |

**Table 3.**
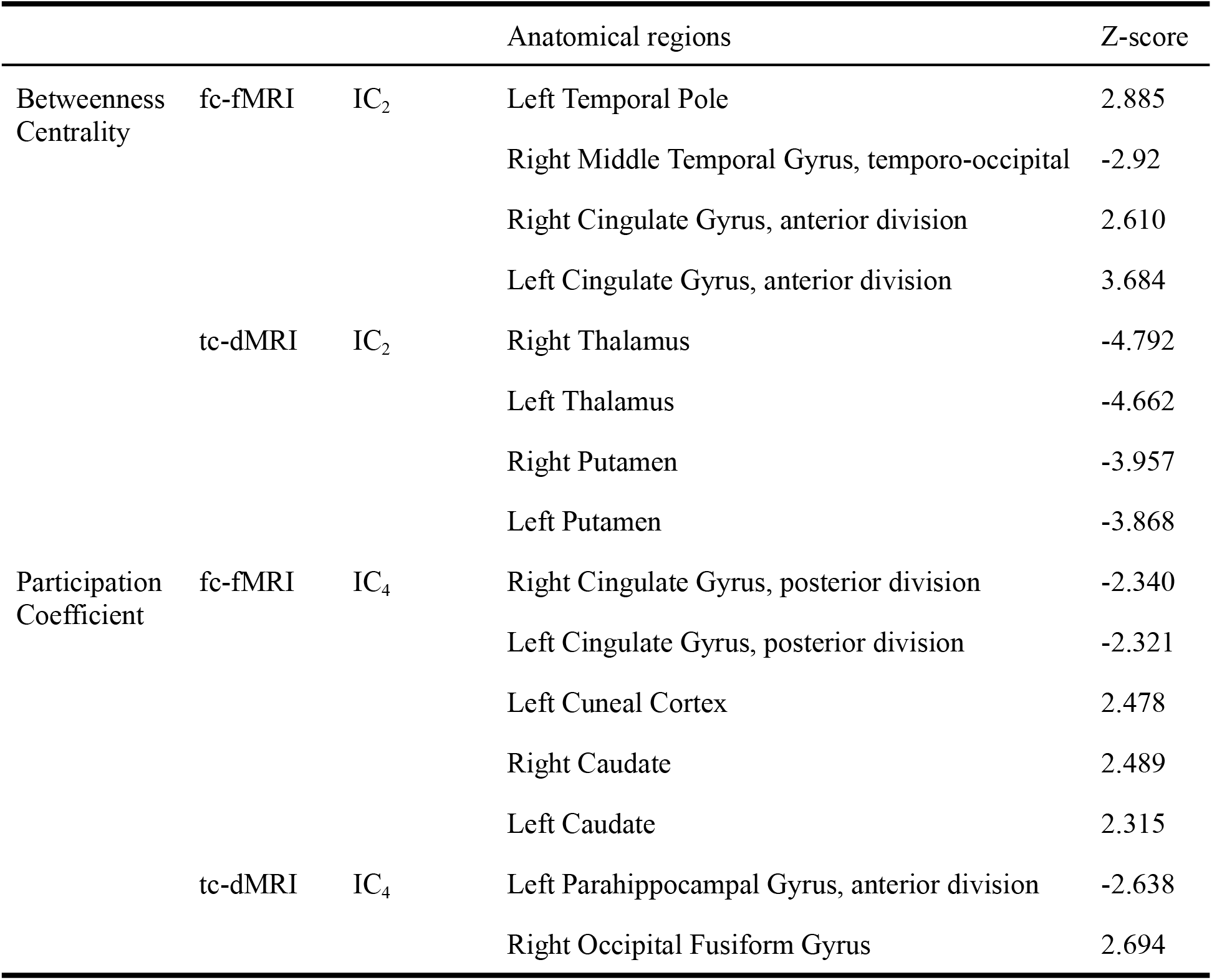
Regions in joint *IC*_*n*_ at *Z* ≥ ± 2. 3

**Figure 4.**
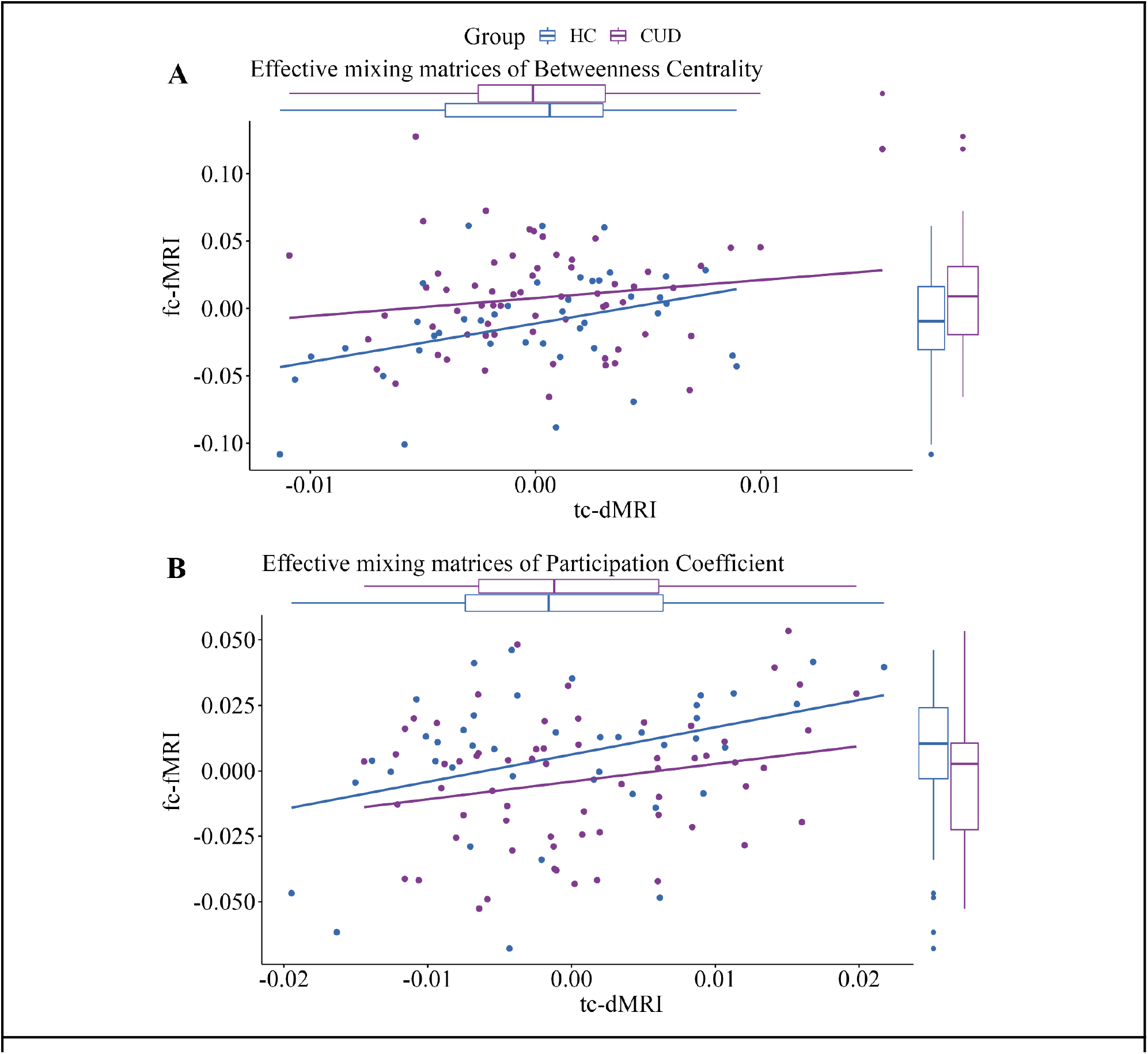
Association between effective mixing matrices of fMRI and dMRI Independent components. A) Association of *emm*_*fc−fMRI*_ *and emm*_*tc−dMRI*_ for betweenness centrality (BC IC_2_) with a significant correlation (r = 0.196, p_fdr_ = 0.045), B) Association of *emm*_*fc−fMRI*_ *and emm*_*tc−dMRI*_ for participation coefficient (PC IC_4_) with a significant correlation (r =0.223, p_fdr_ = 0.044).

## Discussion

In this study, we examined the macroscopic network-based differences using graph theory, between patients with cocaine use disorder (CUD) and matched healthy controls (HC) by combining fc-fMRI and tc-dMRI imaging modalities. While multimodal fusion has been carried out in other studies, to our knowledge, this is the first multimodal-fusion study that uses graph theory to explore topological alterations in CUD patients. First, we evaluated the differences between the two groups in functional and structural modalities separately. Subsequently, we performed a multimodal fusion of fc-fMRI and tc-dMRI modalities, which enabled us to understand network patterns conveyed by both modalities, leveraging a well-defined mathematical framework (i.e. graph theory). While this study was exploratory in nature, post rigorous multiple comparison corrections, our study identified brain regions that not only agrees with earlier studies but also revealed interesting observations that may contribute to a better understanding of CUD patients. We found impairment in inter-module communication (i.e., participation coefficient) in CUD in individual and joint modalities. However, impairment in internode information communication was observed in CUD only in joint modalities. These results demonstrate the utility of multimodal fusion in unearthing latent network patterns which would otherwise be lost if done separately.

Unimodal analysis indicated a reduced contribution of left caudate to inter-modular communication among the CUD when compared to HC in the fc-fMRI modality. The caudate nucleus, a part of the striatum, has been described as a core region involved in habit learning, motor behavior, and compulsive drug-seeking behavior (30). The present findings are in line with previous MRI studies which have found striatum alteration in CUD patients such as a reduced striatal volume (4,31), morphological and microstructural changes (32,33), and altered functional connectivity (34–36). A lower inter-modular communication of the caudate nucleus in CUD may be related to compulsive drug-seeking behavior. Using graph-theory-based multimodal fusion analysis, we found other subcortical regions appearing in addition to the caudate nucleus found in the unimodal analysis. The joint components involved subcortical nodes such as the putamen and thalamus, as well as cerebral cortex such as the anterior/posterior cingulate, parahippocampus, medial temporal, occipital fusiform gyrus, and cuneal cortex, which are commonly associated with CUD (2,4,37).

Brain networks of CUD patients revealed a lesser number of shortest pathways via nodes, reflected by higher betweenness centrality (BC) in fc-fMRI modality (i.e. the contribution rate of nodes in the information interchange with other nodes). This pattern in BC was observed in the Temporo-occipital part of the middle temporal gyrus (TO2) in CUD, a region related to multimodal sensory integration (38) and cognitively implicated in the organization of communicative information (39), emotion recognition, empathic arousal, and retrieval of relevant schemas (i.e. moral judgments) (40). This region has also been shown to be impaired in CUD (33,41) and associated with cue reactivity/craving in cocaine and other SUDs (42–45).

On the other hand, the disruption of the bilateral anterior cingulate gyrus (ACC) in terms of connectivity has been extensively observed in CUD patients (41,46). In recent years, ACC is considered one of the main potential biomarkers and targets for brain stimulation treatments, such as repetitive transcranial magnetic stimulation, due to the strong structural and functional connectivity with the reward system and executive-salience networks (47,48). As reflected by our results, the disturbances in the communication of this region could lead to the reorganization of brain networks observed in CUD (48). Overall, the higher BC observed in certain regions of CUD patients could be indicative of an alteration in the organization of the functional networks. While the unimodal analysis did not reveal these functional changes, the multimodal analysis resulted in identifying these alterations.

Although there have been few studies that have investigated the role of brain regions in inter-network communication particularly, it is noteworthy that we found a lower participation coefficient (PC) in the caudate nucleus among the CUD when compared to HC in both unimodal and multimodal analyses. The reduced participation coefficient displayed by the caudate nucleus along with the posterior cingulate cortex (PCC) (internally oriented processing), an important region of the default mode network, suggests a reduced role they both play in inter-module information transfer in CUD when compared to HC. Previously, Liang et al., (2015) reported a lower PC of both anterior and posterior cingulate cortex in CUD, and connected with regions associated with executive control network (externally oriented executive functioning), explaining the cognitive difficulties in these patients (49). Liang et al., also found a close connection of the caudate nucleus over different brain networks altered in CUD (49,50). Several studies have highlighted both regions as critical hubs vulnerable to cocaine misuse and other SUDs (51–53). One of the main reasons may be the high metabolic costs involved in the process of integration and information exchange within brain networks, both implicated and affected not only in SUD pathology, but in Alzheimer’s-type neurodegeneration, dementia and depression (49).

## Limitations

Despite the findings, there are two main limitations in the present study. The first one is the missed significance in tc-dMRI independent components and the other limitation is related to the lack of correlation with clinical metrics. The no-group differences in the tc-dMRI modality were observed in unimodal or multimodal analyses, which could be attributed to metrics based on graph theory, which are used to detect higher-order relationships in the brain. In both unimodal and multimodal analyses, these higher-order dependencies were manifested as distinct functional differences. Furthermore, this could also explain the lack of correlations with clinical measures. In addition, the existence of a subclinical and/or cognitive profile within the CUD group could also explain the lack of correlations with clinical measures.

## Conclusion

In summary, we found unimodal and multimodal cocaine impairment in inter-module communication and internode exchange communication only in a multimodal manner. Unimodal results show a striatal decrease in the participation coefficient, but the applied supervised data fusion could reveal other regions with cocaine-related impairments in joint-functional communication. Further research applying the combination of modalities is needed to develop better pre-treatment/post-treatment intervention designs and to provide new insights into the neurobiological mechanisms of CUD.

## Supporting information

Supplementary results

## Data Availability

The MRI data is available for download at https://openneuro.org/datasets/ds00334679. Please download the latest available version as there may be updates. Clinical measures are available in Zenodo https://doi.org/10.5281/zenodo.512333180.

https://doi.org/10.5281/zenodo.512333180

## Data and code availability

For the code analysis presented here, please check: https://github.com/psilantrolab/

## Acknowledgements

We thank the people who helped this project in one way or another: Thania Balducci Garcia, Ernesto Reyes Zamorano, Jorge J. Gonzalez Olvera, Francisco J. Pellicer Graham, Margarita López-Titla, Aline Leduc, Erik Morelos-Santana, Diego Angeles Valdez, Alely Valencia, Lya Paas, Daniela Casillas, Sarael Alcauter, Luis Concha and Bernd Foerster. We also thank Rocio Estrada Ordoñez and Isabel Lizarindari Espinosa Luna at the Unidad de Atención Toxicológica Xochimilco for all their help and effort. Finally, we thank the study participants for their cooperation and patience. This study received support from Luis Aguilar, Alejandro De León and Carlos Flores of the Laboratorio Nacional de Visualización Científica Avanzada (LAVIS). Finally, we thank Leopoldo Gonzalez Santos for his technical support. This project was funded by CONACYT-FOSISS project No. 0201493 and CONACYT-Cátedras project No. 2358948. JR-T is a doctoral student from the Programa de Doctorado en Ciencias Biomédicas, Universidad Nacional Autónoma de México (UNAM) and received fellowship 858667 from CONACYT.

## Authors contribution

EAGV conceived the experiments; EAGV and VA, designed the analysis; SS and JR-T analyzed the data; JR-T, SS, EGV and VA wrote the manuscript; EAGV and VA supervised the entire process, and all authors contributed to interpretation of data, revised the manuscript and approved the final manuscript.

## Supplementary method

### Graph measures

Assortativity is the correlation between the degree of a node and the average degree of the node’s neighbors. A positive correlation indicates an assortative network, whereas a negative correlation indicates a disassortative network. Disassortative networks indicate strong hierarchical configurations.

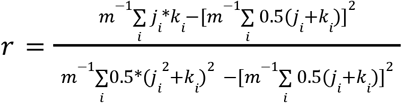

j_i_ and k_i_ represent the degrees of the vertices j and k connecting the i^th^ edge, with i = 1,2, ..m; where m is the total number of edges.

Network efficiency is defined as the average inverse shortest path length in a network. Network efficiency for a network G is as follows.

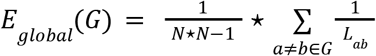

*L*_*ab*_ is the shortest path between nodes a and b in the network G. N is the total number of nodes in the network.

Modularity (M) is a statistic used to distinguish between the number of intra-module connections of an existing network and randomly connected edges in a random network; it tells us how good the clustering is.

Small world index is defined as the ratio of the normalized clustering coefficient and normalized characteristic path length.

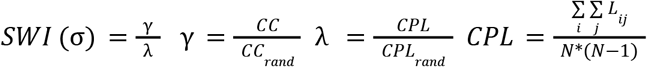

CC and CPL are the clustering coefficient and characteristic path length of the actual brain network, whereas CC_rand_ and CPL_rand_ are generated using 100 random networks by the Markov-chain algorithm. Here L_ij_ is the shortest path between node i and node j.

The hierarchical coefficient (β) quantifies the presence of the hierarchical organization in a network and Synchronization is defined as the ratio of the second smallest Eigenvalue to the largest Eigenvalue obtained through the coupling matrix of a network G.

The Betweenness Centrality of a node (v) is defined as the ratio of the number of shortest paths passing through the node between any two given nodes σ_*ab*_(*v*) to the total number of shortest paths between the two given nodes σ_*ab*_. If the betweenness centrality is high, it means the information flow through that node is high.

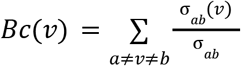

The Degree Centrality of a node(v) is defined as the ratio of the degree of the node (*d*_*v*_) to the maximum possible degree of the node. If the degree centrality of a node is high, it means it is more central.

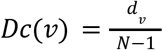

Where N is the total number of nodes in the network.

The Participation coefficient of a node(v) reflects the within-module and intermodular communication.

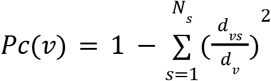

*N*_*s*_ is the number of modules, *d*_*vs*_ is the degree of a node ‘v’ to the nodes in module ‘s’, *d*_*v*_, is the total degree of node ‘ V ’ is 0 when there are no intermodular connections.

Nodal local efficiency is similar to network efficiency, but it is computed in the neighborhood of a node.

The clustering coefficient of a node(v) is defined as the ratio of the number of connections between the neighbors of the node (*e*_*v*_) to the total number of possible connections among (*K*_*v*_) neighbors of the node. If the clustering coefficient of a node is high, it means the neighbors of the node are well connected. It measures the cohesiveness in a network.

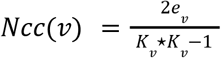

## References

1. Volkow ND, Michaelides M, Baler R (2019): The Neuroscience of Drug Reward and Addiction. Physiol Rev 99: 2115–2140.

2. Lees B, Garcia AM, Debenham J, Kirkland AE, Bryant BE, Mewton L, Squeglia LM (2021): Promising vulnerability markers of substance use and misuse: A review of human neurobehavioral studies. Neuropharmacology 187: 108500.

3. Hanlon CA, Canterberry M (2012): The use of brain imaging to elucidate neural circuit changes in cocaine addiction. Subst Abuse Rehabil 3: 115–128.

4. Pando-Naude V, Toxto S, Fernandez-Lozano S, Parsons CE, Alcauter S, Garza-Villarreal EA (2021): Gray and white matter morphology in substance use disorders: a neuroimaging systematic review and meta-analysis. Transl Psychiatry 11: 29.

5. Costumero V, Rosell Negre P, Bustamante JC, Fuentes-Claramonte P, Adrián-Ventura J, Palomar-García M-Á, et al. (2021): Distance disintegration characterizes node-level topological dysfunctions in cocaine addiction. Addict Biol e13072.

6. Zhang Y, Zhang S, Ide JS, Hu S, Zhornitsky S, Wang W, et al. (2018): Dynamic network dysfunction in cocaine dependence: Graph theoretical metrics and stop signal reaction time. Neuroimage Clin 18: 793–801.

7. Wang Z, Suh J, Li Z, Li Y, Franklin T, O’Brien C, Childress AR (2015): A hyper-connected but less efficient small-world network in the substance-dependent brain. Drug Alcohol Depend 152: 102–108.

8. Sui J, Adali T, Yu Q, Chen J, Calhoun VD (02/2012): A review of multivariate methods for multimodal fusion of brain imaging data. J Neurosci Methods 204: 68–81.

9. Calhoun VD, Sui J (2016): Multimodal Fusion of Brain Imaging Data: A Key to Finding the Missing Link(s) in Complex Mental Illness. Biological Psychiatry: Cognitive Neuroscience and Neuroimaging, vol. 1. pp 230–244.

10. Sui J, He H, Yu Q, Chen J, Rogers J, Pearlson GD, et al. (2013): Combination of Resting State fMRI, DTI, and sMRI Data to Discriminate Schizophrenia by N-way MCCA + jICA. Front Hum Neurosci 7: 235.

11. Oertel-Knöchel V, Reinke B, Feddern R, Knake A, Knöchel C, Prvulovic D, et al. (2014): Episodic memory impairments in bipolar disorder are associated with functional and structural brain changes. Bipolar Disord 16: 830–845.

12. Kim S-G, Jung WH, Kim SN, Jang JH, Kwon JS (2015): Alterations of Gray and White Matter Networks in Patients with Obsessive-Compulsive Disorder: A Multimodal Fusion Analysis of Structural MRI and DTI Using mCCA+jICA. PLoS One 10: e0127118.

13. Meade CS, Li X, Towe SL, Bell RP, Calhoun VD, Sui J (2021): Brain multimodal co-alterations related to delay discounting: a multimodal MRI fusion analysis in persons with and without cocaine use disorder. BMC Neurosci 22: 51.

14. Angeles-Valdez D, Rasgado-Toledo J, Issa-Garcia V, Balducci T, Villicaña V, Valencia A, et al. (2022): The Mexican magnetic resonance imaging dataset of patients with cocaine use disorder: SUDMEX CONN. Scientific Data, vol. 9. https://doi.org/10.1038/s41597-022-01251-3

15. Paliwal P, Hyman SM, Sinha R (2008): Craving predicts time to cocaine relapse: further validation of the Now and Brief versions of the cocaine craving questionnaire. Drug Alcohol Depend 93: 252–259.

16. World Health Organization (2010): Measuring Health and Disability: Manual for WHO Disability Assessment Schedule WHODAS 2.0. World Health Organization. Retrieved from https://books.google.com/books/about/Measuring_Health_and_Disability.html?hl=&id=h9fhLNiaRTgC

17. McLellan AT, Thomas McLellan A, Kushner H, Metzger D, Peters R, Smith I, et al. (1992): The fifth edition of the addiction severity index. Journal of Substance Abuse Treatment, vol. 9. pp 199–213.

18. Patton JH, Stanford MS, Barratt ES (1995): Factor structure of the barratt impulsiveness scale. Journal of Clinical Psychology, vol. 51. pp 768–774.

19. Esteban O, Markiewicz CJ, Blair RW, Moodie CA, Isik AI, Erramuzpe A, et al. (2019): fMRIPrep: a robust preprocessing pipeline for functional MRI. Nat Methods 16: 111–116.

20. Ciric R, Rosen AFG, Erus G, Cieslak M, Adebimpe A, Cook PA, et al. (2018): Mitigating head motion artifact in functional connectivity MRI. Nat Protoc 13: 2801–2826.

21. Power JD, Mitra A, Laumann TO, Snyder AZ, Schlaggar BL, Petersen SE (2014): Methods to detect, characterize, and remove motion artifact in resting state fMRI. Neuroimage 84: 320–341.

22. Tournier J-D, Smith R, Raffelt D, Tabbara R, Dhollander T, Pietsch M, et al. (2019): MRtrix3: A fast, flexible and open software framework for medical image processing and visualisation. Neuroimage 202: 116137.

23. Wang J, Wang X, Xia M, Liao X, Evans A, He Y (2015): GRETNA: a graph theoretical network analysis toolbox for imaging connectomics. Front Hum Neurosci 9: 386.

24. Danon L, Duch J, Arenas A, Díaz-Guilera A (2007): Community Structure Identification. Large Scale Structure and Dynamics of Complex Networks. pp 93–114.

25. Rubinov M, Sporns O (2010): Complex network measures of brain connectivity: uses and interpretations. Neuroimage 52: 1059–1069.

26. Li Y-O, Adalı T, Calhoun VD (2007): Estimating the number of independent components for functional magnetic resonance imaging data. Human Brain Mapping, vol. 28. pp 1251–1266.

27. Himberg J, Hyvarinen A (n.d.): Icasso: software for investigating the reliability of ICA estimates by clustering and visualization. 2003 IEEE XIII Workshop on Neural Networks for Signal Processing (IEEE Cat. No.03TH8718). https://doi.org/10.1109/nnsp.2003.1318025

28. Abrol A, Rashid B, Rachakonda S, Damaraju E, Calhoun VD (2017): Schizophrenia Shows Disrupted Links between Brain Volume and Dynamic Functional Connectivity. Front Neurosci 11: 624.

29. Revell AY, Silva AB, Arnold TC, Stein JM, Das SR, Shinohara RT, et al. (2022): A framework For brain atlases: Lessons from seizure dynamics. Neuroimage 254: 118986.

30. Lipton DM, Gonzales BJ, Citri A (2019): Dorsal Striatal Circuits for Habits, Compulsions and Addictions. Front Syst Neurosci 13: 28.

31. Barrós-Loscertales A, Garavan H, Bustamante JC, Ventura-Campos N, Llopis JJ, Belloch V, et al. (2011): Reduced striatal volume in cocaine-dependent patients. Neuroimage 56: 1021–1026.

32. Garza-Villarreal EA, Chakravarty MM, Hansen B, Eskildsen SF, Devenyi GA, Castillo-Padilla D, et al. (2017): The effect of crack cocaine addiction and age on the microstructure and morphology of the human striatum and thalamus using shape analysis and fast diffusion kurtosis imaging. Transl Psychiatry 7: e1122.

33. Rasgado-Toledo J, Shah A, Ingalhalikar M, Garza-Villarreal EA (2022): Neurite orientation dispersion and density imaging in cocaine use disorder. Prog Neuropsychopharmacol Biol Psychiatry 113: 110474.

34. Zhang S, Li C-SR (2018): Ventral striatal dysfunction in cocaine dependence – difference mapping for subregional resting state functional connectivity. Transl Psychiatry 8: 1–11.

35. Zhang S, Wang W, Zhornitsky S, Li C-SR (2018): Resting State Functional Connectivity of the Lateral and Medial Hypothalamus in Cocaine Dependence: An Exploratory Study. Front Psychiatry 9: 344.

36. Hu Y, Salmeron BJ, Gu H, Stein EA, Yang Y (2015): Impaired functional connectivity within and between frontostriatal circuits and its association with compulsive drug use and trait impulsivity in cocaine addiction. JAMA Psychiatry 72: 584–592.

37. Poireau M, Milpied T, Maillard A, Delmaire C, Volle E, Bellivier F, et al. (2022): Biomarkers of Relapse in Cocaine Use Disorder: A Narrative Review. Brain Sci 12. https://doi.org/10.3390/brainsci12081013

38. Mesulam MM (1998): From sensation to cognition. Brain 121 (Pt 6): 1013–1052.

39. Papeo L, Agostini B, Lingnau A (2019): The Large-Scale Organization of Gestures and Words in the Middle Temporal Gyrus. J Neurosci 39: 5966–5974.

40. Garrigan B, Adlam ALR, Langdon PE (2016): The neural correlates of moral decision-making: A systematic review and meta-analysis of moral evaluations and response decision judgements. Brain Cogn 108: 88–97.

41. Camchong J, MacDonald AW 3rd, Nelson B, Bell C, Mueller BA, Specker S, Lim KO (2011): Frontal hyperconnectivity related to discounting and reversal learning in cocaine subjects. Biol Psychiatry 69: 1117–1123.

42. Courtney KE, Schacht JP, Hutchison K, Roche DJO, Ray LA (2016): Neural substrates of cue reactivity: association with treatment outcomes and relapse. Addict Biol 21: 3–22.

43. Tapert SF, Cheung EH, Brown GG, Frank LR, Paulus MP, Schweinsburg AD, et al. (2003): Neural response to alcohol stimuli in adolescents with alcohol use disorder. Arch Gen Psychiatry 60: 727–735.

44. van Hell HH, Vink M, Ossewaarde L, Jager G, Kahn RS, Ramsey NF (2010): Chronic effects of cannabis use on the human reward system: an fMRI study. Eur Neuropsychopharmacol 20: 153–163.

45. Witteman J, Post H, Tarvainen M, de Bruijn A, Perna EDSF, Ramaekers JG, Wiers RW (2015): Cue reactivity and its relation to craving and relapse in alcohol dependence: a combined laboratory and field study. Psychopharmacology 232: 3685–3696.

46. Spronk DB, van Wel JHP, Ramaekers JG, Verkes RJ (2013): Characterizing the cognitive effects of cocaine: a comprehensive review. Neurosci Biobehav Rev 37: 1838–1859.

47. Verdejo-Garcia A (2018): New Insights on Neurocognition in Cocaine Use Disorder. Current Behavioral Neuroscience Reports 5: 232–237.

48. Zhao Y, Sallie SN, Cui H, Zeng N, Du J, Yuan T, et al. (2020): Anterior Cingulate Cortex in Addiction: New Insights for Neuromodulation. Neuromodulation. https://doi.org/10.1111/ner.13291

49. Liang X, He Y, Salmeron BJ, Gu H, Stein EA, Yang Y (2015): Interactions between the salience and default-mode networks are disrupted in cocaine addiction. J Neurosci 35: 8081–8090.

50. Menon V (2011): Large-scale brain networks and psychopathology: a unifying triple network model. Trends Cogn Sci 15: 483–506.

51. Tomasi D, Volkow ND, Wang R, Carrillo JH, Maloney T, Alia-Klein N, et al. (2010): Disrupted functional connectivity with dopaminergic midbrain in cocaine abusers. PLoS One 5: e10815.

52. Ma L, Steinberg JL, Moeller FG, Johns SE, Narayana PA (2015): Effect of cocaine dependence on brain connections: clinical implications. Expert Rev Neurother 15: 1307–1319.

53. Zhang R, Volkow ND (2019): Brain default-mode network dysfunction in addiction. Neuroimage 200: 313–331.

