## Supplementary results for "Structural and functional pathology in cocaine use disorder with polysubstance use: a multimodal fusion approach"

### Supplementary figures

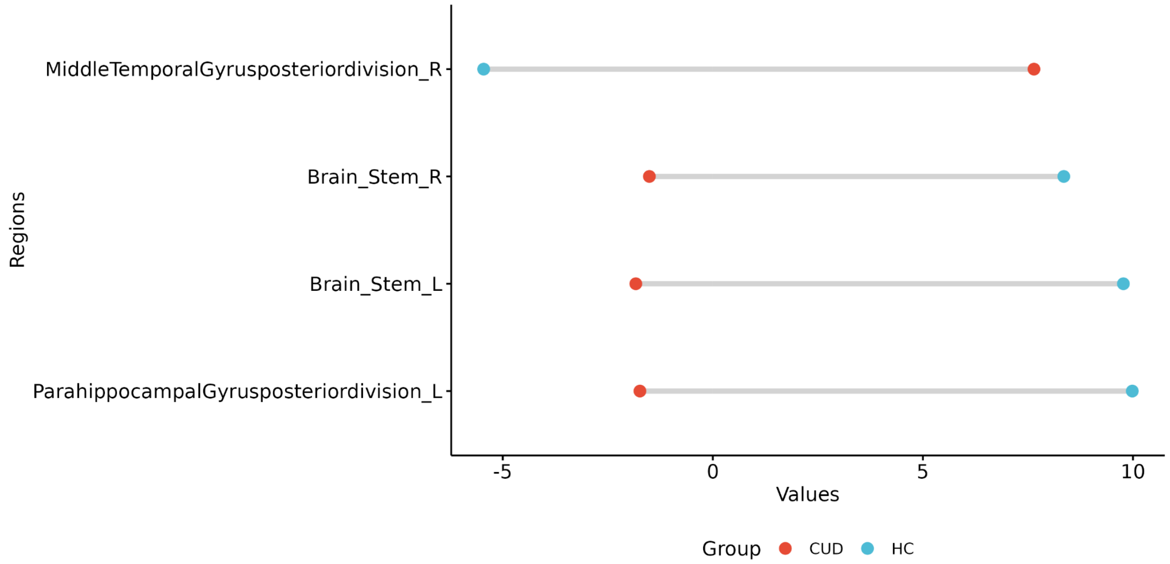


SFigure 1. Group comparison between cocaine use disorder (CUD) vs healthy controls (HC) in betweenness centrality (BC) using Harvad-Oxford atlas at unimodal sMRI approach. Values are uncorrected by multiple comparisons.

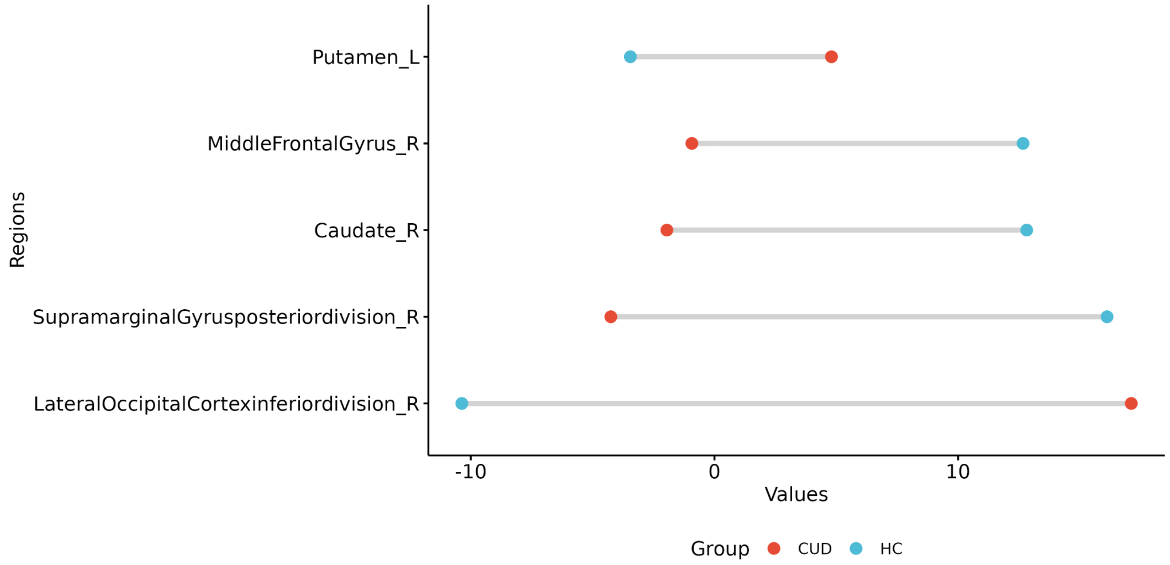


SFigure 2. Group comparison between cocaine use disorder (CUD) vs healthy controls (HC) in betweenness centrality (BC) using Harvad-Oxford atlas at unimodal fMRI approach.

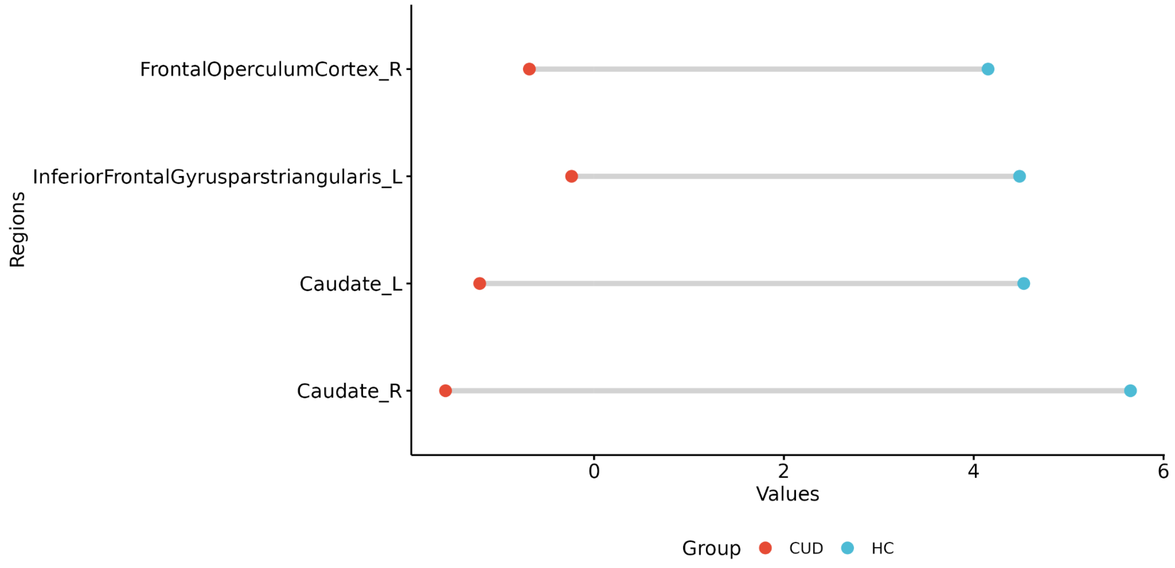


SFigure 3. Group comparison between cocaine use disorder (CUD) vs healthy controls (HC) in degree centrality (DC) using Harvad-Oxford atlas at unimodal fMRI approach. Values are uncorrected by multiple comparisons.

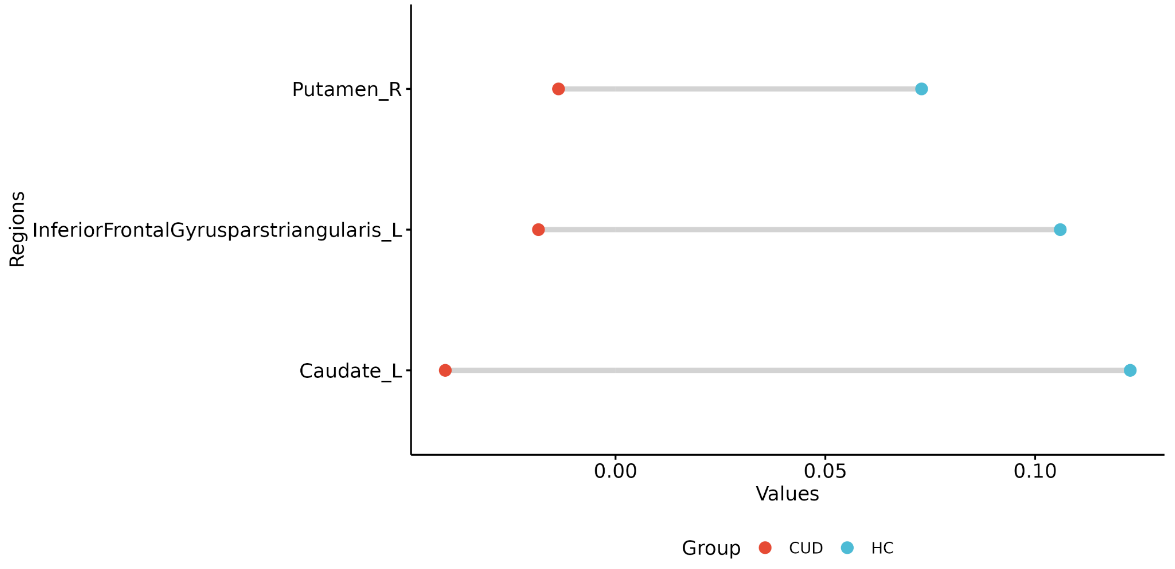


SFigure 4. Group comparison between cocaine use disorder (CUD) vs healthy controls (HC) in participation coefficient (PC) using Harvad-Oxford atlas at unimodal fMRI approach. Values are uncorrected by multiple comparisons.

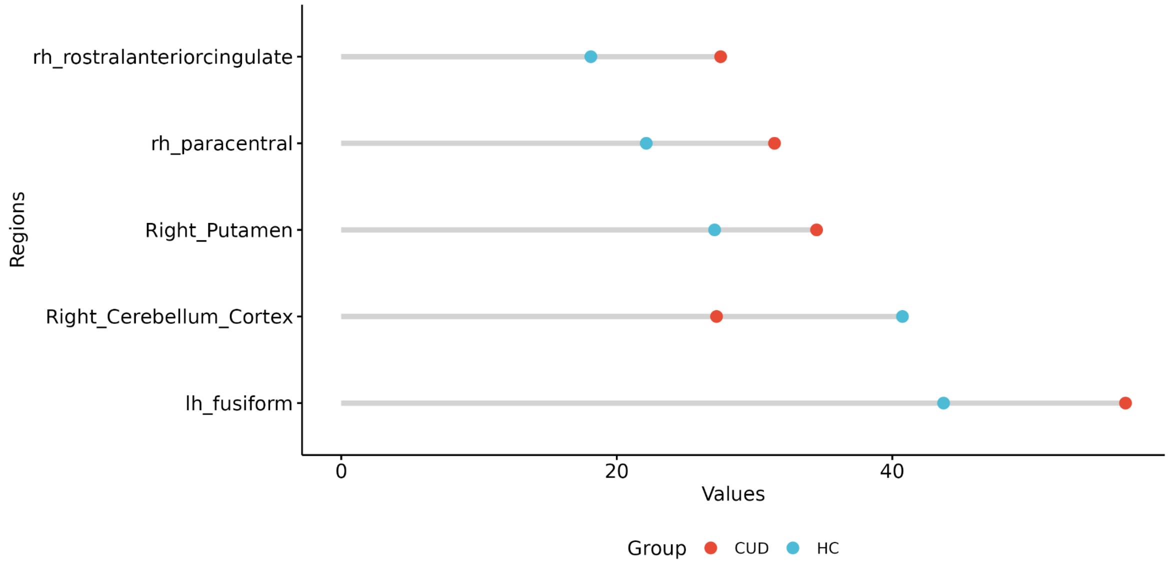


SFigure 5. Group comparison between cocaine use disorder (CUD) vs healthy controls (HC) in betweenness centrality (BC) using Desikan-Killiany atlas at unimodal fMRI approach. Values are uncorrected by multiple comparisons.

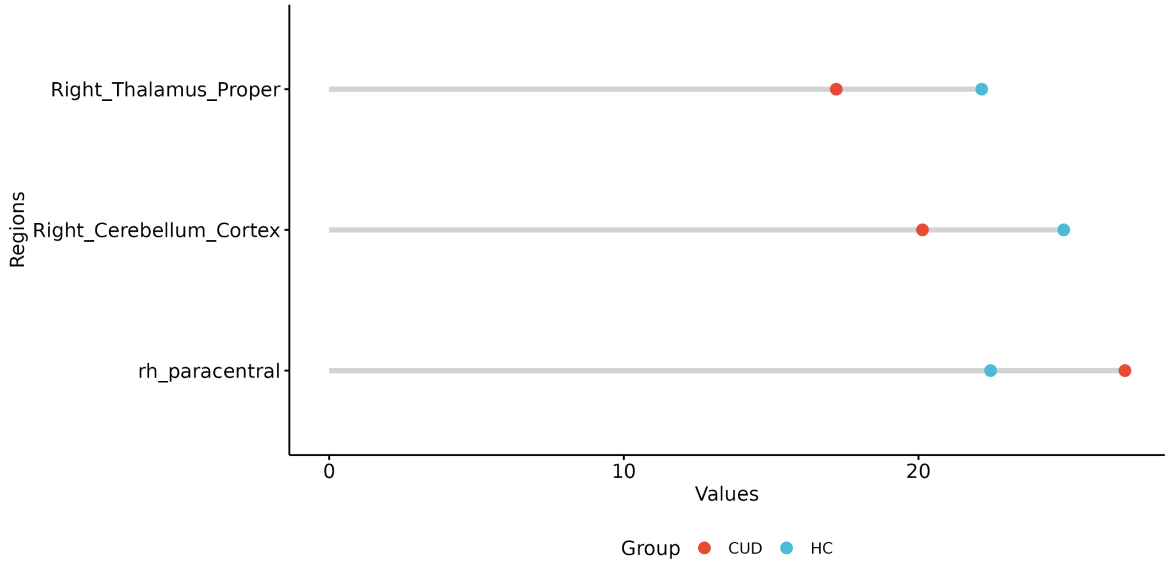


SFigure 6. Group comparison between cocaine use disorder (CUD) vs healthy controls (HC) in degree centrality (DC) using Desikan-Killiany atlas at unimodal fMRI approach. Values are uncorrected by multiple comparisons.

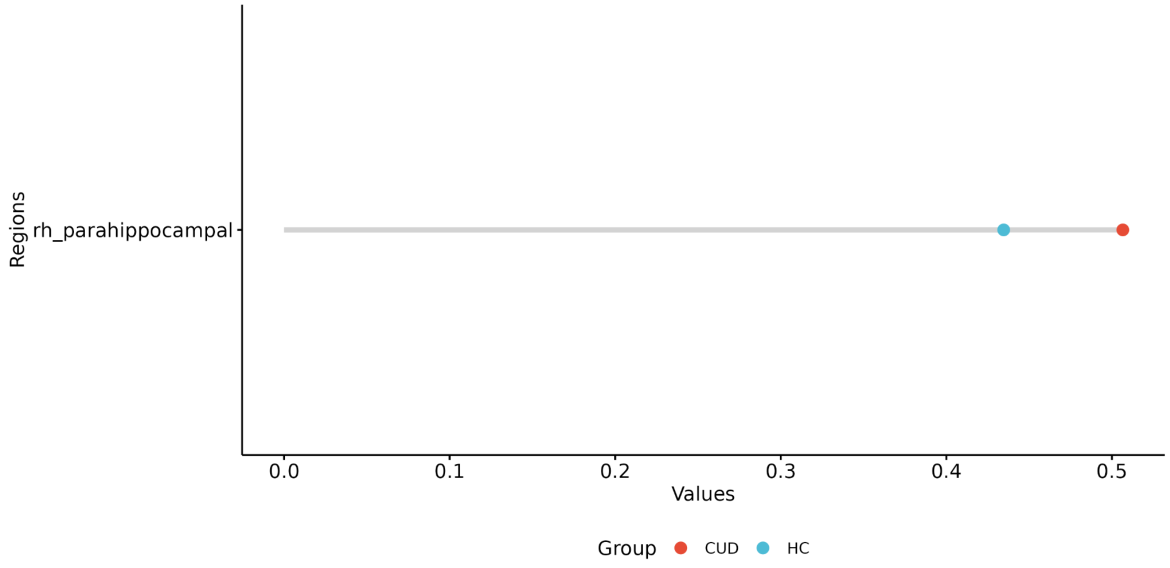


SFigure 7. Group comparison between cocaine use disorder (CUD) vs healthy controls (HC) in nodal clustering coefficient (NCC) using Desikan-Killiany atlas at unimodal fMRI approach. Values are uncorrected by multiple comparisons.

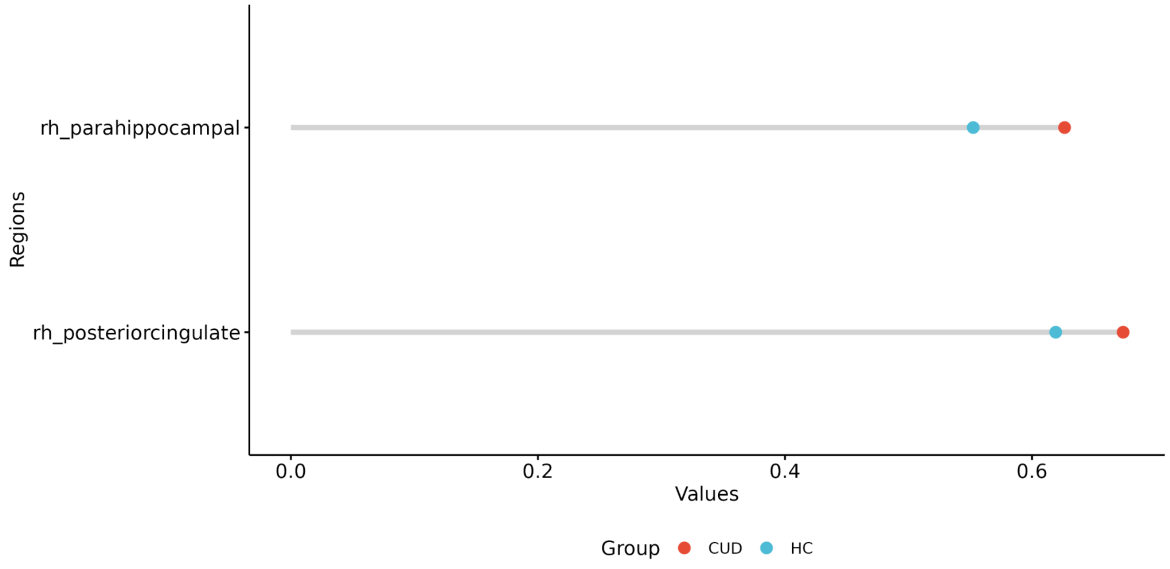


SFigure 8. Group comparison between cocaine use disorder (CUD) vs healthy controls (HC) in nodal local efficiency (NLE) using Desikan-Killiany atlas at unimodal fMRI approach. Values are uncorrected by multiple comparisons.

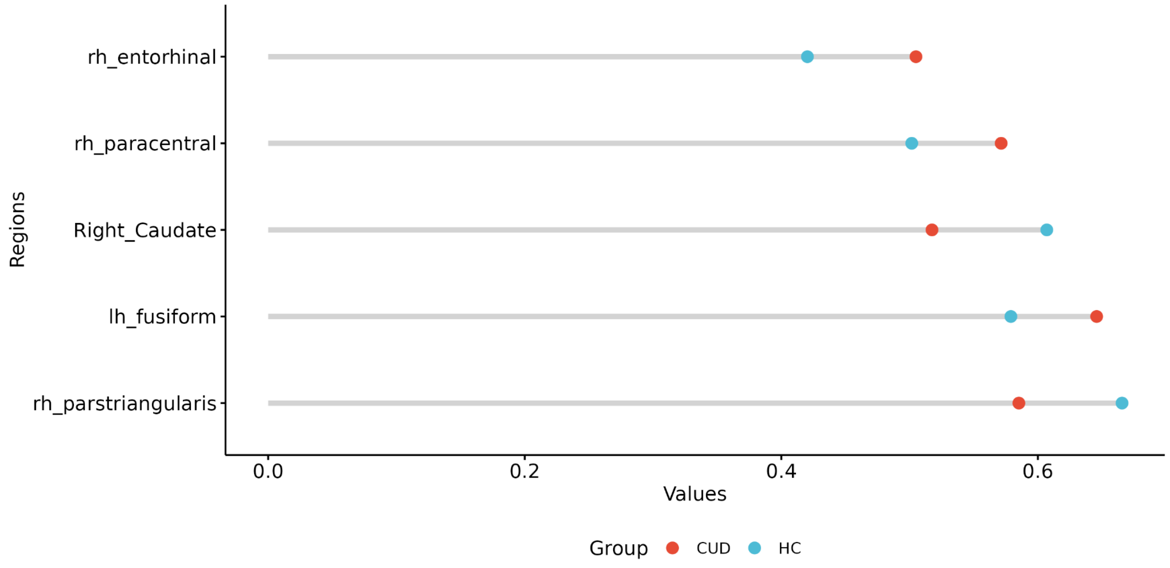


SFigure 9. Group comparison between cocaine use disorder (CUD) vs healthy controls (HC) in participation coefficient (PC) using Desikan-Killiany atlas at unimodal fMRI approach. Values are uncorrected by multiple comparisons.
